# A fair efficacy formula for assessing the effectiveness of contact tracing applications

**DOI:** 10.1101/2020.11.07.20227447

**Authors:** Adam Fowler

## Abstract

Mobile contact tracing apps have been developed by many countries in response to the COVID-19 pandemic. Trials have focussed on unobserved population trials or staged scenarios aimed to simulate real life. No efficacy measure has been developed that assesses the fundamental ability of any proximity detection protocol to accurately detect, measure, and therefore assess the epidemiological risk that a mobile phone owner has been placed at. This paper provides a fair efficacy formula that can be applied to any mobile contact tracing app, using any technology, allowing it’s likely epidemiological effectiveness to be assessed. This paper defines such a formula and provides results for several simulated protocols as well as one real life protocol tested according to the standard methodology set out in this paper. The results presented show that protocols that use time windows greater than 30 seconds or that bucket their distance analogue (E.g. RSSI for Bluetooth) provide poor estimates of risk, showing an efficacy rating of less than 6%. The fair efficacy formula is shown in this paper to be able to be used to calculate the ‘Efficacy of contact tracing’ variable value as used in two papers on using mobile applications for contact tracing [6]. The output from the formulae in this paper, therefore, can be used to directly assess the impact of technology on the spread of a disease outbreak. This formula can be used by nations developing contact tracing applications to assess the efficacy of their applications. This will allow them to reassure their populations and increase the uptake of contact tracing mobile apps, hopefully having an effect on slowing the spread of COVID-19 and future epidemics.

## 1 Introduction

Many teams worldwide have developed contact tracing applications. Many of them use Bluetooth Low Energy (BLE) technology present in most mobile phones. In future it is envisaged that new sensors and communication protocols will augment or replace BLE for contact tracing. Examples could include Ultra Wide Band (UWB) radio or ultrasound based protocols. Also envisaged is the use of wearable devices specific for contact tracing. In order for the contact tracing applications to be useful in stopping the spread of an infection like COVID-19 their Bluetooth Proximity Detection Protocol must accurately identify and measure the distance between phones running the same application. If a particular protocol under records then it will under record risk, leading to fewer notifications of exposure than is needed, hindering control of virus spread. If a protocol over estimates risk then too many people will be notified unnecessarily leading to a loss of faith in mobile contact tracing, followed by reduced compliance to its notifications, and again less control of virus spread. Recent papers have concentrated on calculating risk scores from distance estimation values using RSSI values and the time & duration of a contact event [3]. This paper defines the probability of detection of the risk that occurs within a contact event, and how closely it maps to the actual risk incurred by an individual. A Contact Event is the time and distance of two people within close proximity, at least to a distance of likely exposure to infection. As two people approach each other, meet, and then part their risk exposure will vary according to time spent at each distance from each other. Contact tracing applications, and the proximity detection protocols within them, aim to accurately measure this exposure risk. When one party to the contact event falls ill they can share their contact events such that those placed at high risk of infection are notified and can self isolate, limiting the spread of a disease in a population.

There are many areas a protocol can fail to accurately measure risk. Firstly if a particular contact event is not recorded at all by not supporting a particular phone, or scanning for nearby contacts too infrequently. Closely following a contact event by regularly estimating distance, which we call continuity, is where risk can be over or under estimated. Not regularly measuring duration with distance around the riskiest (closest) part of a contact event - having poor completeness of a risk profile - can also lead to poor recording of exposure risk. Similarly not recording the distance accurately enough can lead to errors in risk calculation. An app must also have the longevity to work throughout a normal working day in order to provide the most useful service. Finally, the population reach of a protocol - how many people can use it given their mobile phone’s capabilities - will limit the control effect of any protocol. These are the constituent measures we define in this paper that make up our fair efficacy formula. These issues are shown in Figure 1, below.

**Fig. 1.**
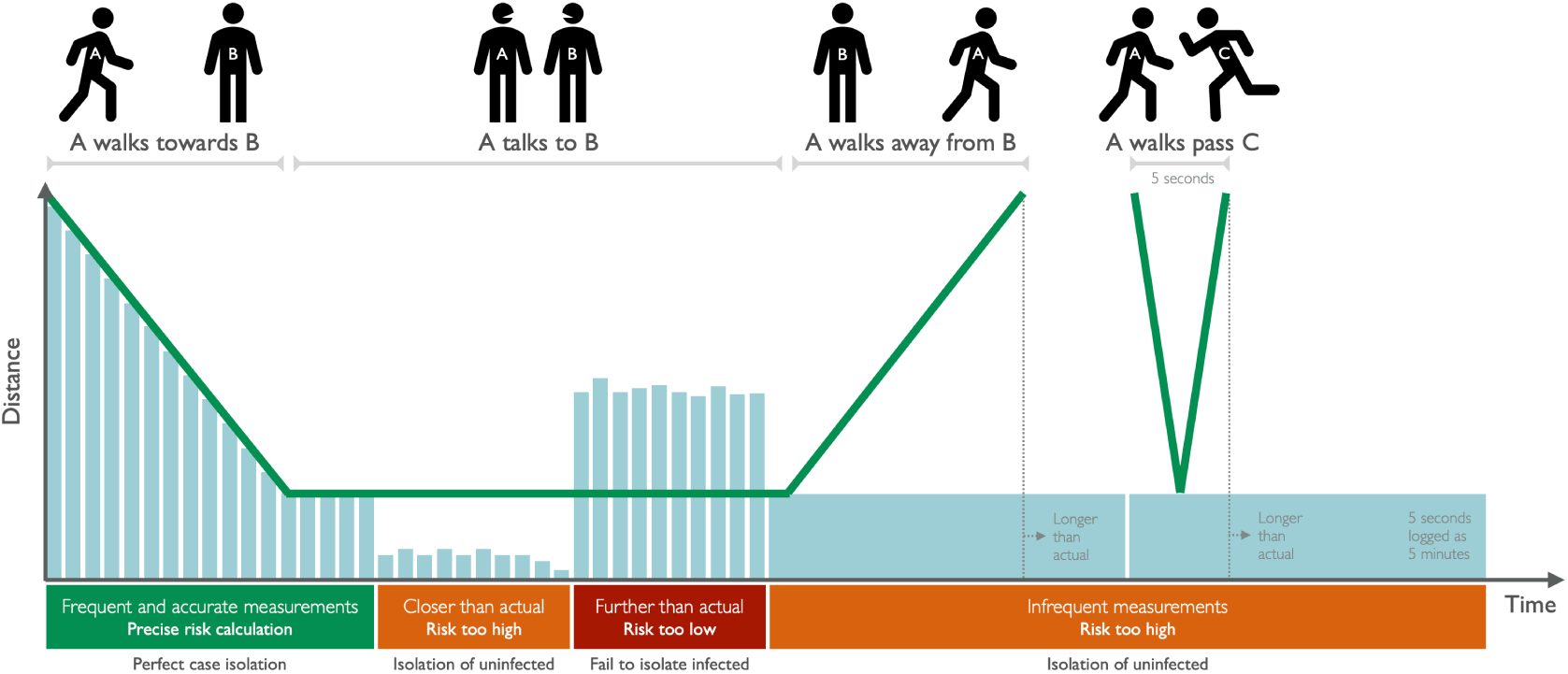
How over and under estimation of risk exposure can happen

This paper uses an example contact event with synthetic data to illustrate concepts such as time windows, distance estimations, and different ways a protocol may measure time, and duration. Some protocols may, for example, only give the time of a contact event to an approximate time window. Others may bucket their distance analogue (E.g. For radio based protocols like Bluetooth this would be the Received Signal Strength Indicator - RSSI) in to bands. Bracketing exposure time or bucketing distance analogue introduces limitations for such protocols’ accuracy at measuring a contact event. This will result in either over or under estimating an individual’s exposure and chance of infection. This results in lower epidemiological efficacy, limiting protocols’ ability to slow the spread of a disease. We highlight these limitations in this paper, and provide a fair efficacy formula for measuring a protocol’s effectiveness.

Our primary contributions in this paper are: - A novel protocol for measuring the efficacy of contact tracing apps - A detailed comparison of existing contact tracing approaches using our formula - A new contact tracing protocol that achieves over 86% probability of accurate risk detection, resulting in 38% efficacy

## 2 Related work

The original work on mathematically modeling of the control effect of an instant contact tracing application was published by Ferretti et al in the journal Science. [6] They discuss the standard 3 days to find and isolate a contact with manual contact tracing and show that such a delay results in a health system’s inability to control the spread of COVID-19 under any circumstances. The paper goes on to say “Traditional manual contact-tracing procedures are not fast enough for SARS-CoV-2. However, a delay between confirming a case and finding that person’s contacts is not inevitable. Specifically, this delay can be avoided by using a mobile phone app.” This analysis goes on to plot success in isolating patients against the ability to effectively trace their contacts instantly using a mobile application as we describe in this paper. That analysis (Fig 3 in the paper) shows that a range of efficacy - from 90% effective instant contact tracing with 10% isolation of patients, to 60% effective instant contact tracing with 90% isolation of patients, will go beyond the margin of error for R0 measurement and have a controlling effect on the spread of COVID-19. A protocol will need 30% efficacy in order to be effective. Below 10% a protocol will not be effective

It should be noted that the above paper talks about ‘detection’ in the context of ‘accurate risk detection’ rather than ‘did two phones see each other’. These are very different concepts. Accurate risk detection has epidemiological usefulness, device detection alone does not. My paper will consider ‘detection’ to mean ‘accurate risk detection’. As a result, the ‘Efficacy’ score in the Oxford paper is what we term ‘Probability of risk detection’ in this paper.

It should be noted that these figures apply only to first degree contacts. I.e. direct contacts. If a so-called *centralised model* is used then second and third degree contacts could also be traced [8], providing more control over spread than *decentralised models* that only allow first degree contacts to be traced. It should also be noted that the efficacy percentage quoted in the quoted paper consists of the following elements “The efficacy of contact tracing (the y axis of Fig. 3) is the square of the proportion of the population using the app, multiplied by the probability of the app detecting infectious contacts, multiplied by the fractional reduction in infectiousness resulting from being notified as a contact.”. Our paper provides a standardised way to measure and define the probability of the app detecting exposure risk. In this paper we shall use the as a basis of analysis as this is where the author lives.

The Ferretti et al paper [6] builds on earlier work from Fraser et al [7] that modeled ways an infectious disease outbreak could be controlled. In particular this provided the Tracing effectiveness measure *ε*_*T*_. This comprises the product of the square of population reach, the probability of detecting exposure risk, and a fixed factor of how much effect on reducing disease spread contact tracing has. The paper does not provide a mechanism for defining population reach or probability of detection.

Our paper provides a way of measuring proximity detection protocols and calculating their *ε*_*T*_ value using a standard set of tests that allow protocols to be directly compared to one another in terms of their epidemiological efficacy.

In the UK The Alan Turing Institute and the University of Oxford’s Fraser Group have worked on the theory behind much of the epidemiological need for contact tracing applications. They have produced a range of papers on risk scoring calculations [3], the control effect of test-trace-isolate on the Isle of Wight [5], and on processing raw RSSI values from contact tracing apps [1].

These papers and others discuss what to do with the indicators from these mobile apps after data collection has been completed. The inputs for the algorithms and approaches suggested in these papers relies upon accurate and timely data being generated from contact tracing applications and their underlying proximity detection protocols and their contact event data recordings. There is a need to measure the reliability of the generation of this initial data in order to avoid a ‘garbage in, garbage out’ problem where the data input to the later estimation and risk scoring algorithms is such that the reliability of their outputs, and thus the efficacy of control on a pandemic, cannot be relied upon.

We continue in this paper to discuss ways the reliability of this input data can be measured, and thus how contact tracing application protocols can be compared. We do not discuss converting this raw data in to distance estimations, or risk models based on this data, as these have been discussed in previous papers. [2]

## 3 Applying the fair efficacy formula

We also show that the formula can be applied to a real proximity detection protocol and its code. We have used the formula to assess and direct development of a new proximity detection protocol called Herald. [18] Over the course of 5 weeks of development we improved its efficacy score from 13% to over 40% in formal testing. We are providing this code under an open source license along with its analysis R scripts in order to assist worldwide teams in improving their own protocols.

This application of our fair efficacy formula on a real protocol shows the utility of a comprehensive efficacy measure and standard tests in order to rapidly improve a protocol. These proximity detection protocols are a fundamental part of contact tracing applications. It is the hope of the author that national contact tracing teams apply the formula to their own Bluetooth protocols, helping to improve public health outcomes during the COVID-19 pandemic.

## 4 Efficacy indicators

A variety of descriptions of efficacy for proximity detection protocols have been discussed. Researchers from The Alan Turing Institute have suggested [19] the following three broad categories:-

1. Continuity - The ability of a proximity detection protocol to take a set of detections and correctly identify a single real world contact event throughout the entire contact event period
2. Completeness - At and around the moment in the encounter of maximum risk, and thus minimum separation distance, how complete is the recorded dataset and how closely does it fit the actual distance
3. Accuracy - How accurate is the data produced that is later passed to a risk scoring algorithm. This is a function of the accuracy of the distance between individuals, or a modified annealed distance estimation of raw data, such as Received Signal Strength Indicator (RSSI)

In addition to these in order to provide enough information of epidemiological usefulness to a national health response, and in order to ensure continued reliability during a full working day, we shall add the following categories too:-

1. Detection - Whether at any point during a contact event a device saw another device (one-way detection), or whether both saw each other (two-way detection). This is included separately to continuity as if a phone isn’t detected its presence will not show up in continuity data, giving a false sense of success.
2. Population Reach - Given the prevalence of Mobile Phones in a particular country and those phones’ capabilities, this is the maximal proportion of people that can use a protocol
3. Longevity - The ability of an application to continue to detect and range a set of contact events throughout a working day, or whether a protocol in practice degrades during a day

This paper suggests measures and tests for all of the above efficacy indicators. This paper then combines these measures to come up with a way to empirically measure the contact tracing efficacy variable *ε*_*T*_ as defined in the 2004 Fraser et al paper. [7]

An often overlooked component of the data provided by the proximity detection protocols is their correction, bucketing, or other processing of the raw Bluetooth data before a description of a contact event is passed to a contact tracing application. This processing can be a mean averaging over time, or grouping distances from continuous measures in to distance bands, or buckets. These changes can result in artificially high or low risk exposure as recorded by some protocols. These issues are discussed under the Continuity heading as they are often related to detection time delay and time annealing - both functions of continuity. We also discuss distance bucketing under Accuracy.

## 5 Contact event definition

Before we proceed there is a need to standardise our definitions of terminology, independent of specific protocols. There is a long list of terminology which is used in common parlance amongst those working on contact tracing apps and proximity protocols. We provide a standard glossary of definitions on our website. [17]

Before we can determine measures it is first necessary to understand a real world timeline for a contact event (Actual contact event) versus what is recorded by a proximity detection protocol (Measured contact event) and passed to a contact tracing application.

We shall consider a single contact event between two phones, A and B, with the minimum separate phases that occur over time.

1. Two devices are operable and running the same protocol but are not yet within detection range of each other. This is phase 0 (P0)
2. As the two phone owners approach each other phone A is detected by B, but phone B is not yet detected by A. This is phase 1 (P1) and occurs at t1 in the contact event. Recorded as RSSI1. This situation is generally described as ‘one way detection’.
3. Closer still, phone A is both detected by B, but also detects B itself. This is phase 2 (P2). This situation is called ‘two way detection’.
4. The two people enter the range of interest to epidemiologists because of disease transmission. This is phase 3 (P3)
5. The two people and devices are at the nearest distance *D*_*NEAREST*_ to each other for this encounter. This is phase 4 (P4)
6. The two people start to move apart but are still in the range of interest to epidemiologists (as at P3). This is phase 5 (P5)
7. Getting further apart, phone A is detected by phone B, but phone B is not detected by phone A (as at P1). This is phase 6 (P6). Again this is ‘one way detection’.
8. The two devices exit detection range, and the contact event ends (as at P0). This is phase 7 (P7) and occurs at *t*_*N*_ in the contact event. The last event measurement is recorded as *RSSI*_*N-*1_, but the contact event ends at time *t*_*N*_ (which is *t*_*N-*1_ + *FREQ*_*DETECTION*_ for the particular protocol)

The above phases are illustrated in the below diagram:-

## 6 Measures of continuity

Continuity is the ability of a protocol to detect and range two devices during their actual physical contact event. We define minimum acceptance criteria and a set of *measures* for each aspect of continuity.

### 6.1 Minimum requirements for providing continuity

It is first desirable to state the minimum effective criteria of a proximity detection protocol. For a valid epidemiological recording of a contact event to occur, P3 must occur after P2 and P1. Likewise, P5 must occur before P6 and P7. I.e. the detection must occur before the devices enter the range of epidemiological interest, and not end until they leave this range.

If a given protocol records P1 and P2 as happening after P3 has been actually reached then it cannot be said *to provide continuity coverage* across the whole range of time of a contact event. The same is true if P4 occurs after P5 and P6.

If the protocol records P2 only as happening after P3, or P4 occurring after only P5, then it is said *to provide partial continuity coverage* across the whole range of time of a contact event.

If neither of the above is true, then the protocol cannot be said to provide continuity and thus is said *to not provide continuity coverage* across the whole range of time of a contact event.

The above is a discrete measure with three possible values, and so a formal mathematical definition is not provided.

The remaining measures discussed in this paper only provide a basis of comparison between protocols if both protocols provide continuity coverage across the whole range of time of a contact event. If that is not possible then the protocol that cannot provide continuity coverage is the inferior one as it may not detect a contact event at all.

It’s important to note that continuity is not just a question of range and transceiver power. If a protocol has a large time detection window then it may not detect a short lived but risky contact event at all, or mis-attribute time and risk within such an event, as described in the example below.

#### 6.1.1 Range of epidemiological interest

There is ongoing research in to the maximum distance at which exposure can be said to have been accrued. WHO guidelines have emphasised the ‘two metre’ rule, but have also given advice about being at ‘one metre with precautions’. A recent paper in the BMJ [4] has suggested we should think of exposure in a more fluid environment, up to 8 metres. They recommend that “Instead of single, fixed physical distance rules, we propose graded recommendations that better reflect the multiple factors that combine to determine risk.” They go on to detail a review of current studies and state “Yet eight of the 10 studies in a recent systematic review showed horizontal projection of respiratory droplets beyond 2 m for particles up to 60 µm. In one study, droplet spread was detected over 6-8 m (fig 2). These results suggest that SARS-CoV-2 could spread beyond 1-2 m in a concentrated packet through coughs or sneezes.” … “Physical distancing rules would be most effective if they reflected graded levels of risk.”. In this paper we use a continuous distance variable as the desired outcome, and therefore presume a continuous variable as the distance analogue (E.g. RSSI for Bluetooth). We also calculate the impact that a protocol using ‘buckets’ for its distance analogue would have on the accuracy, and thus efficacy, of such a proximity detection protocol.

**Fig. 2.**
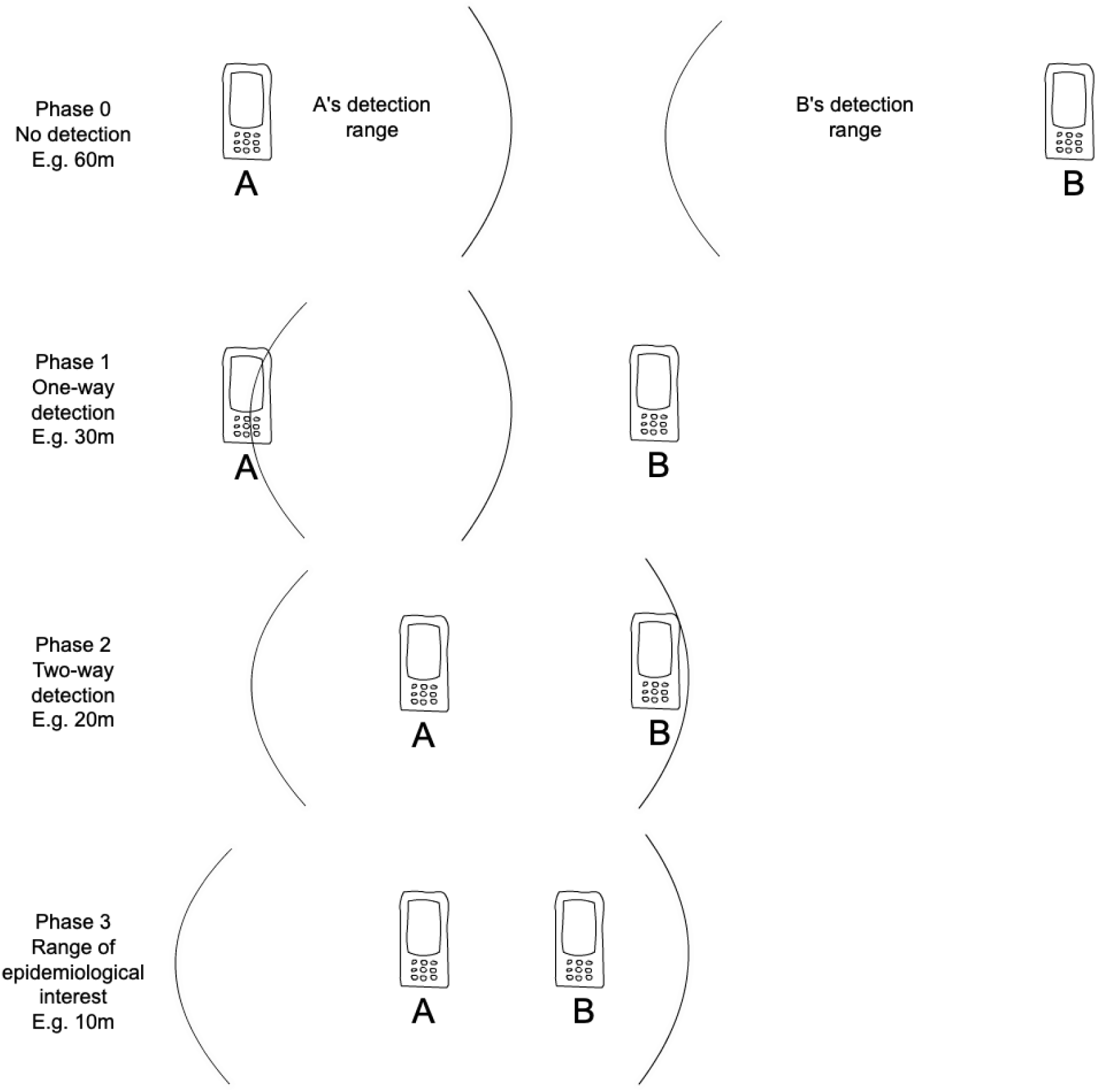
Stages of a Contact Event as measured by a mobile phone

### 6.2 Summary information on a contact event

The following definitions are useful to understand a contact event but do not provide a basis of comparison. The total duration of a single contact event is thus defined as *DUR*_*CE*_ = *t*_*N*_ *-t*_1_ More accurately, the actual time the contact event lasted for is 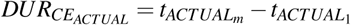 and the time recorded for the contact event by the protocol under analysis is 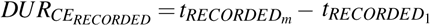

### 6.3 Continuous measures of continuity for comparison

We can specify the difference between the recorded start and end of the contact event and that recorded by the protocol under test as:-

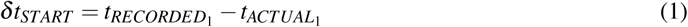

and

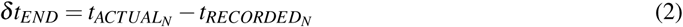

respectively.

These are both measures of the start and end (i.e. maximum extent) of continuity. As they represent a fixed number of seconds they do not need to be scaled like other measures - they already provide a directly comparable measure between protocols without scaling.

In an ideal algorithm both delta values should approach zero compared with what has actually happened, thus:-

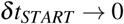

and

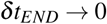

If a given protocol exhibits time windows that exceed the contact event start and end times then the above two measures will be negative. This will be an indication of over-recording (and thus over risk estimation) for that protocol.

The further from zero either of these numbers are, the less accurately the risk exposure accrued to an individual is measured, potentially leading to delayed or missed exposure notification (a false negative), or risk notification sent out when less risk was actually accrued (a false positive).

#### 6.3.1 Detection time windows

Whilst clock skew on devices is an issue, the use of time synchronization services means this risk is minimised, and so such an accurate windowing time period is possible. Similarly, mobile phone operating systems’ own bluetooth protocol stack have a timeout and will cut a bluetooth connection if a response to a request (A BLE characteristic read, write or notify) is not received promptly. On Apple’s iOS 13.5.1 this timeout is 30 seconds.

The Oxford Risk Model [6] gives a fixed risk value for contacts at up to 1m in range, and logarithmically falls off with distance after this. In this model the risk required to hit ‘15 minutes at two metres’ will be equivalent to a 1 metre contact of 3.5 minutes duration. Given this fact it is necessary for the time window chosen to be a fraction of this time in order to not over estimate a person’s risk of exposure. The Oxford Risk Model scores all contacts up to a metre in distance the same, so this time period is the shortest possible to create a single ‘dangerous contact’ with an infected individual that will exceed a notifiable level of risk.

We therefore recommend a fair continuity measurement time slice of 30 seconds throughout for formal testing. This paper is written with a maximum detection window size of 30 seconds being acceptable. Some protocols may have more frequent detections. It is possible for a particular protocol to have multiple detections and ranging within a single 30 second window. To enable a fair comparison we restrict comparisons to whether a detection happened or not in each 30 second period. For testing this should be at the start of the minute on the receiving device, and 30 seconds after this, in order to provide a consistent definition local to each device. The smaller this time interval the more accurate a protocol will be in risk estimation.

### 6.4 Common problems limiting continuity measurements, or masking of problems

There are a range of problems we have encountered during testing of our own new protocol that skew risk measurements and make some scenarios look artificially better or worse for than is actually the truth for risk accrual. In this section we describe many of these issues, and approaches that can be taken to eliminate them from comparisons of different protocols.

#### 6.4.1 Under and over estimating risk

Whilst an actual physical detection may happen at a specific point in time and at a specific distance between devices, it is possible that a protocol provides modified, summarised, or otherwise annealed numbers rather than the raw time data to a contact tracing application. In epidemiological terms this introduces some negative side effects in terms of data capture for continuity measurements in the form of both under and over estimating risk during a contact event. These result from the use of time periods rather than continuous readings. The larger the time window between distance analogue measurements, the greater the possibility for under and over recording.

#### 6.4.2 Over and under recording

It is necessary therefore to discuss the differences between the periodic detections in two different theoretical protocols in order to determine a model for continuity comparison measures. Consider the following two protocols, one with a 5 minute detection scan, annealed to a 5 minute block from the top of the hour (i.e. rounded down to a 5 minute block) versus a 30 second average detection scan in a protocol with exact time measurement to the second and no time annealing:-

Along the X axis is the reading number for a simple contact event as discussed earlier in this section (P1 to P7). Annotated on to this axis are the actual points in time for the phases discussed earlier, *t*_*P*1_ to *t*_*P*7_

Along the Y axis is the distance analogue between phones. For a particular protocol this may be a different measure (E.g. RSSI) used as a distance analogue, but here we are considering the general physical event and so use actual distance. The point of nearest distance *D*_*NEAREST*_, and thus maximum risk, is shown on the chart as the lowest red bar. The distance of minimum continuity coverage for the epidemiological needs is *D*_*MIN*_.

#### 6.4.3 Affect of over and under recording on risk

The Alan Turing Institute’s risk paper [3] includes the definition of a function, marked as equation (1) in the paper, for calculation of the total risk score of an encounter. Assuming the environment is consistent throughout the contact event, the only variable quantity during the contact event is distance. The distance function in the Turing risk paper, equation (2), was applied to the above data. This shall be called the risk contribution in this section. This gives the risk accrued during each window as below:-

The raw synthetic data used for these illustrations can be found on our website. [14] The total risk score accrued by the 5 minute annealed recording approach was 6.112. The total risk contribution for the 30 second window, non-annealed, version was 8.046. This is a significant difference. As can be seen by the chart this would have been an even bigger difference in some situations, but in our scenario over recording before the riskiest part of the contact event due to annealed time masks the effect on the accuracy of the risk contribution. I.e. the cancelling out of a large time window makes this sample data look better (i.e. nearer risk scores) than may happen in real life.

The contact event risk score threshold, beyond which an individual would be given advice to take a healthcare action, was calculated for a distance contribution of 2 meters at 15 minutes, giving a risk contribution of 3.75. During our contact event of 20 minutes the risk score threshold would have been reached by either recording method. This means that for longer contact events at a short distance you are less likely to suffer a false negative - i.e. not alerting a person to take action when they have, in reality, exceeded the risk threshold. At longer distances though the chance of a false negative increases.

For shorter contact event durations, however, a five minute window would clearly result in increased incidences of both false negative (under estimation of total risk contribution) and false positive calls to action to users of a contact tracing app, and incorrect information on the number of potentially infected people being passed to national health authorities. For contact event durations of less than the protocol’s average recording window duration there is a high degree of chance of false positives. Consider the situation where you are walking to work and passing people walking the other way. The contact event duration where you are at a distance likely to give you a higher risk contribution would be less than 30 seconds. For a protocol with a five minute detection window it is highly unlikely a contact would be recorded, but if it were it would be recorded for a duration of five minutes. If this point in time coincides with your closest distance of, say, 1 metre, then you only need to walk passed a single ill person for a few seconds to gain a risk score above the 3.75 risk threshold. 4 windows of 30 seconds gives a risk contribution of 4.0, exceeding our threshold after just 2 minutes.

Protocols with time windows close to or exceeding 2 minutes are therefore very likely to result in false positive notifications. This could affect the public’s trust in an app and the likelihood of them complying with a recommendation to self isolate, as well as give a false impression of outbreak progress to national health authorities.

#### 6.4.4 Reduction of control effect due to false indications in large time window protocols

We recommend that as part of any comparison of proximity detection protocols that a short lived set of tests is carried out in order to confirm the designed recording window.

We further recommend that an epidemiological impact assessment or simulation be carried out in a separate study to this paper for the negative effects of any protocol with a recording period greater than 30 seconds, and how that could impact a health authority’s ability to observe and control an outbreak. It is possible that a 5 minute annealed window will lead to so many false positive calls to action to users before the R rate is under control during an outbreak that the number of people being asked to take corrective action such as tests and self isolation is so great as to, effectively, result in incorrectly locking down a large proportion of the population. This could lead not only to a poor control effect in the case of false negatives (missed contact events less than a 5 minute window), but also to a loss of faith in, and thus compliance with, calls to action in the case of too many false positive notifications.

#### 6.4.5 Continuity and radio interference

Detections happen at a particular point in time with the scans that result in these detections (and thus distance estimations) occurring no more often than a maximum scan frequency determined by the protocol itself. For the Herald protocol this has been measured at up to every 4 seconds. The risk accrual is based both on the distance estimation at this point in time multiplied by the time difference between this and the next detection, assumed to be a static distance for this window of time. At worst then this time is the same as the scan interval - 4 seconds for the Herald. Generally, as the average actual mean detection time window size approaches zero seconds, the accuracy increases to its best possible value. Thus the mean and variance of detection periods are useful indicators of potential risk estimation accuracy.

Radio protocols have to deal with the physical reality of the universe and thus are built with interference, intermittent or dropped connections, and data receipt errors in mind. It is perfectly possible, for example, for multiple detection windows to be missed but to maintain a single contact event. This does give problems for accuracy (as you’d be assuming the distance estimation was the same during the ‘missing’ data interval) but not for continuity necessarily. The longer the gap between readings is, the less likely the readings represent the same contact event.

### 6.5 Measuring continuity coverage

For a given contact event with detections at time points t1 to tN there must therefore be at least 1 detection (distance estimation) in each 30 second ‘bucket’ during a contact event attributed to a specific pairing of devices to achieve the best continuity measure. The difference in time between readings must also be presented to the risk scoring algorithm.

It is possible for a detection protocol to have a very ‘lumpy’, or unevenly distributed, set of detections over time. This can occur when there are more devices nearby than can be detected or connected to (depending on protocol) at the same time. Thus calculating a simple mean average time between detections is insufficient to provide a good measure of continuity.

A time period from 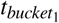 to 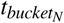 will have 30 seconds between buckets.

A bucket score of 1 at time m is thus defined as:-

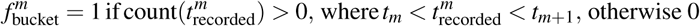

And the scaled continuity rate can thus be calculated as:-

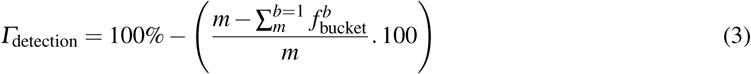

where *m* is the number of 30 second bucket time slices used for comparison. This is multiplied by 100 in the above formula so it appears as a ‘missed bucket percentage’, which is then subtracted from the ideal 100%.

With an ideal algorithm providing a continuity rate approaching 100%:-

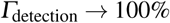

### 6.6 Relative comparisons

If protocol A had a missed bucket percentage of 20% then this protocol could be said to miss the risk during 20% of a contact event. This is why this measure is used in the calculation of the probability of risk detection in the fair efficacy formula.

## 7 Measures of completeness

Completeness aims to discuss the ‘riskiest’ part of a contact event and a protocol’s recorded description of this physical time period. We start by again considering our 5-minute-annealed and 30 second windowed protocols as set out in the Continuity section, above. Here though we concentrate on the area either side of the point of highest risk at *D*_*NEAREST*_, but only to the extent of the risk threshold being reached in the accrued risk score. This leaves us with this data section:-

*D*_*NEAREST*_ is achieved in three consecutive time windows, shown on the above chart at window indexes 7-9. Taken together the risk for these distances does not reach the threshold value (3.7) and so we extend the line by enough buckets for us to exceed this risk score, choosing the adjacent bucket with the next highest risk (lowest distance). For the above data this includes just one additional bucket, bucket 10. These data items taken together are *δ RT*_1_, shown as the area under the red (riskiest part of the encounter) line.

By following the same method we can calculate the *δ RT*_2_ - the readings required to exceed the risk threshold a second time. This is shown by the readings under the orange line.

In a method logically similar to a standard deviation from the centre point of a normal distribution we can therefore define *δ RT*_1_ and *δ RT*_2_

### 7.1 Minimum completeness requirement

There is no minimum boolean completeness requirement as there is for continuity. It is worth nothing, however, that there must be a 30 second window distance estimation at or near the point of highest likely exposure in order for a completeness score to be accurate at all. Imagine, for example, if the red bar risks were missed entirely from a protocol’s record of a contact event. Such a protocol could result in far too little exposure risk being accrued over time during a short contact event. Protocols with a large (>30 seconds) detection window are likely to suffer worst in completeness, and thus epidemiological usefulness, in such a situation as that described in the reference contact event above.

### 7.2 Measuring completeness

We can define completeness - or accurate recording of the riskiest part of the contact event - by reference to the difference in risk accrual as calculated from recorded distance values vs. the actual distance values. For comparison of protocols we shall use the analogue for distance, normally recorded RSSI detection values for Bluetooth based protocols.

To enable this comparison we use a calibration application on two phones placed at a known distance and left for a large period of times, taking thousands of readings. The modal reading is the one that, if accurate, a contact tracing app would be expected to report - *RSSI*_*EXPECTED*_. During actual testing we place the app at a known distance for a short period of time, one minute, and see what RSSI reading was actually recorded - *RSSIRECORDED*

Consider a single reading at *t*_8_ above, with an RSSI of *RSSI*_8_. The accuracy of the 5 minute annealed protocol (purple columns) vs the minimum target epidemiological measurement model (green columns) can be stated as:-

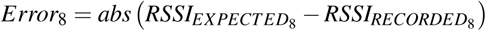

More generically this can be expressed as:-

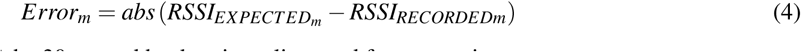

where *m* is the index of the 30 second bucket time slice used for comparison. More generically, and not specific to RSSI on Bluetooth, we can express this as:-

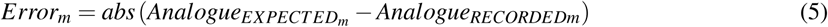

where analogue is the value provided by a given protocol as an analogue to distance (Normally *RSSI* or *attenuation* value).

See 8.2 for a discussion on distance analogue variables

As these individual errors could be numbers in a variety of scales it is necessary to scale them to a known scale before proceeding with analysis. This allows different protocols to be compared to one another. As *absolute* (*RSSI*) provides values between 1 and 99 (with 0 being not in range) we shall use this percentage-like scale for our comparisons to ease the maths for all radio based protocols, thus:-

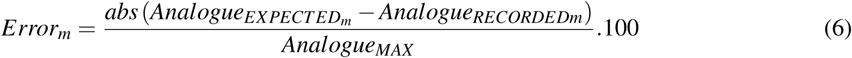

where *m* is the reading number in the sequence within the contact event, and *Analogue*_*MAX*_ is the maximum analogue value in the range of the analogue variable

It is desirable to use the absolute error for each individual time window so errors do not cancel each other out over time, resulting in a lower error score than would occur in real life.

It is assumed that Analogue range begins at 1 (far away, but within detection range) and *Analogue*_*MAX*_ (devices touching or extremely close). If a particular analogue is not based on a value of 1, or uses an inverted number for range, correct it before use in the above formulae.

A negative value for this Error measure will represent an over estimation of RSSI and thus over estimation of risk. More likely, a positive value will represent an under estimation of risk at this time index. The higher the difference in values the worse the fit.

Considering the entire period gives the scaled completeness rate for RT1 as follows:-

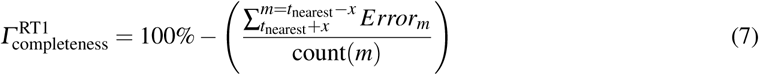

where *x* is the minimum number of buckets either side of the point of minimum distance (maximum RSSI strength and risk) sufficient to achieve an accrued risk above the risk threshold. As this may be such a small number of windows depending on the distance of the contact event (a minimum of four 30-second windows at a range of 1 meter given the Oxford Risk Model, as above), it may be necessary to use the next most significant set of time windows to compare the completeness of two different algorithms.

Thus we should also quote the completeness for the next set of buckets required to achieve the risk threshold - not including the risk buckets used in the previous calculation (as they would skew the result). This gives a scaled completeness rate for RT2 as follows:-

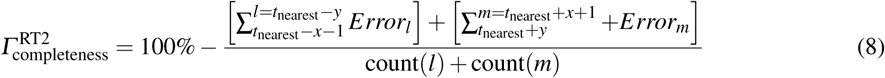

where *y* is the number of buckets either side of the point of minimum distance (maximum RSSI strength and risk) sufficient to achieve an accrued risk of twice the risk threshold.

In an ideal protocol with ideal fitness of the measured distance analogue (RSSI for Bluetooth) we would expect these measures to tend to zero, thus:-

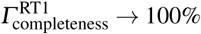

and

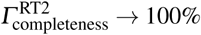

The formal calculation on efficacy only uses the RT1 error value as this is the one under the Oxford Risk Model whose error will most adversely affect the usefulness of a proximity detection protocol for the purposes of epidemiological control of an outbreak.

### 7.3 Short or less risky contact events

Given the Oxford Risk Model is based on accumulated risk we must also consider protocol performance tests where the whole contact event accrues risk but not sufficiently so to breach the risk threshold in this single event. This could happen if, for example, two people are sat somewhat close to each other in a dining area, or pass in the street.

We must come up with a measure for completeness for field tests of a shorter duration or larger distance in order to allow comparisons for completeness in such scenarios.

In such a scenario we can presume a controlled environment whereby different protocols are put through the same tests, with the same phones in the same positions for the same durations. In such a case the total fit of the risk profile in its entirety for the whole contact event can be compared.

By adapting 7 above, we can specify a scaled completeness rate:-

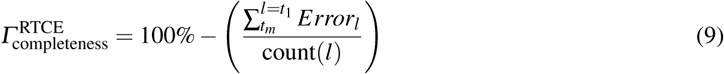

where *t*_1_ and *t*_*m*_ are the times of the start and end of the contact events, respectively. RTCE means Risk Threshold sum for the whole Contact Event.

Again, in an ideal protocol with perfect fit, this value shall tend to 100%:-

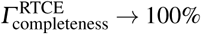

### 7.4 Bucket-wise measuring

It should be noted that the above comparisons are for a 30 second bucket, or time window, only - not the protocol’s own bucket period. This is in order to ensure a consistent comparable measure across protocols for all measure types in this paper. Thus a large error for a single window in a protocol with a large (>30 seconds) recording window will accrue more error overall, as this error will be counted for each 30 second part of that detection window. This is true to the effect of that error on the accrued risk in reality, and so a fair measure for our purposes in this paper.

## 8 Measures of accuracy

The accuracy of a given protocol relates to its ability to correctly measure and calculate the distance at a given point in time between devices. Some protools render a raw RSSI value from Bluetooth, others a mean, modal, or otherwise processed RSSI value, and others still their own analog for distance and RSSI. The GAEN protocol [12] is an example of this latter approach as it provides *attenuation values* rather than RSSI readings.

Whichever value is presented by the protocol to the consuming contact tracing application its effect on the subsequent accuracy of the risk scoring mechanism must be evaluated in order to determine the potential efficacy of a proximity detection protocol.

It should also be noted that the less well a protocol scores against the measures of continuity and completeness the less accurate it will be. You cannot have accurate data if you do not record the data at the appropriate point in time. As these aspects are discussed elsewhere in this paper we shall not repeat their effect in our accuracy analysis.

We must consider a proximity detection scenario in order to determine what measures of accuracy make sense in addition to those mentioned above.

In order to measure accuracy one must deploy a protocol on a pair of phones in a known environment and at a known distance. The other measures in this paper deal with the life of a contact event and how their measures affect overall total risk score accuracy over time. We shall instead now consider a static pair of phones located at a fixed distance over a period of time.

The reason for a longer time period at each distance is to ensure that we mitigate the risk of accidentally measuring the nearest distance exactly accurately as a fluke of recording. This is a possibility given the variability of RSSI at a fixed distance and the use of integers rather than continuous numbers to represent RSSI values.

### 8.1 Prior practical observations

There are a variety of factors that affect the accuracy of a distance estimation analogue. These are out of scope of this paper, but we have summarised our own findings on our research website. [13]

### 8.2 Properties of an accurate distance analogue variable

We consider the analogue to distance as the field whose accuracy needs to be assessed. RSSI is typical. It should be noted that in Bluetooth (and other radio) chipsets RSSI is not a continuous variable but rather a signed integer with values ranging from −1 (weak signal) to −99 (strong signal). The choice of values is not standardised and is left to the particular chipset manufacturer. This is why calibration data is necessary in order to create an accurate distance estimation function based on RSSI values.

Of particular interest is the recent finding by this author that some phones use a distance-logarithm scale for their RSSI recordings whereas others use an inverse distance square based scale for their RSSI readings. Apple iPhone 7 and 7+ use a log-distance scale, whereas the iPhone 6 and iPhone X pairing have been observed to use an inverse square of distance scale. This leads to inaccuracy of distance data around the 2m and above the 5m marks. This means that calibration data and formulae used to convert distance analogues (RSSI) to distance values cannot use a single estimation formula - multiple formulae and distance calibration data points must be used to accurately calibrate phone estimation data.

Other protocols, such as GAEN [12], provide an *attenuation* value which is similarly albeit less finely bucketed than raw RSSI values.

Whichever approach is taken the variable provided by a given protocol as an analogue for distance can be considered as a number on a continuum.

The value provided should have the following properties:-

1. For a known distance and accurate distance estimation function, the variable should provide a value with a small (ideally zero) error from the expected analogue value at that known distance. This relates to both movement and static scenarios. This is the concept of *fit* and has been considered in the Continuity section earlier in this paper. By measuring Continuity across different tests, both static scenarios and those involving constant movement, continuity provides a useful indication of potential accuracy too.
2. If a set of readings are taken in order to produce the analogue value then they should be taken as close together as possible and not spread out across the detection window. This will minimise error due to movement. Thus a set of readings can either be *singular, rapid* or *distributed* within the detection window. Thus *rapid* estimations are likely to be more accurate across contact events involving large amounts of movement. (Restaurant, walking down the street, through a bar, shopping centre, and so on)
3. With devices at a known fixed distance, the variation of values provided across multiple detection windows should be low, ideally with zero variation. This shall be considered below.
4. The time between detections should be as small as possible, but at most the period needed to make an effective epidemiological assessment of risk exposure. As previously mentioned, this is taken as 30 seconds in this paper. This shall be discussed in the section on window time measurement, below.

### 8.3 Continuous versus bounded ranges

It should be noted that any processing, including restricting the distance analogue to particular buckets, will have an adverse effect on accuracy. As an example, if an analogue value of 800 was recorded at, for example, 2 metres but a value of 1200 was recorded at 1 metre, it would be logical to expect the measurement at 1.5m to be 1000. If the particular protocol only bounds its analogue values in bands of 400 though, this would be recorded as 800 or 1200, incurring an error of 200, a significant amount. It is important, therefore, to conduct accuracy measurements at a set of epidemiologically relevant distances.

Given the Oxford Risk Model has maximum risk at or below 1m, and the difference in risk from 1m to 1.1m reduces by 16%, we recommend distance accuracy be calculated for an overall set of distances of 0.8m, 1.0m, 1.2m, 1.5m, 2.0m, 2.5m, 3.0m. I.e. intervals from 0.8m to 3.0m as these are those most important to risk scoring. No weight shall be given to the accuracy score based on relative risk as this is the purpose of the risk estimation function and out of the scope of this paper. I.e. it would be distance estimation function dependent and thus discounted in our calculations.

### 8.4 Measuring accuracy

As previously discussed we shall define value variation and window time intervals as measurements for accuracy.

#### 8.4.1 Distance analogue value accuracy

As discussed in equation 6 above, the error at a particular point in time can be stated as below:-

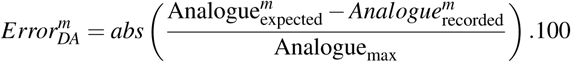

Note: For RSSI based protocols, *abs* (*Analogue*_*MAX*_) and the multiple of 100 cancel each other out to 1. We now have a sequence of analogue errors for a known fixed distance across a set of buckets 1…m. We can now consider this a standard sequence from which we can produce standard mathematical mean and variance calculations.

For an ideal algorithm the outliers in this sequence should tend toward the ideal error value, which is of course zero, thus:-

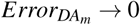

and

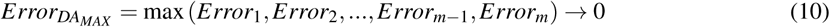

and

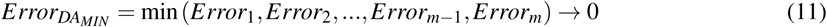

The mean shall also tend this way, and because the mean of error is centered around zero, the variance shall also tend this way, thus:-

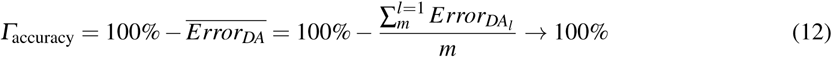

where m is the number of total readings in the contact event at this fixed distance And thus variance can be expressed as:-

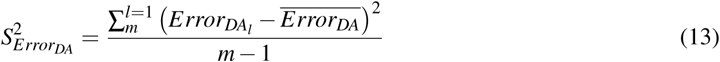

And again it follows that in an ideal protocol the variance should tend to zero too, along with min, max, and mean, thus:-

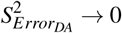

In conclusion the measures we shall use for accuracy are primarily equations 12 and 13, as variance cannot be relied upon alone. Secondarily, to provide more detail on the reason for a higher than desired variance and mean error we recommend that 10 and 11 are also considered during protocol development.

#### 8.4.2 Window time intervals and accuracy

Let us consider an ideal protocol. This protocol will be able to determine exact distance estimation at any point in time down to the nanosecond or smaller. Practically this is not possible due to physical limitations of our universe and the technologies humanity has so far developed. So we must consider what an approximation to the ideal would operate like.

Let us name the as-designed time window as:-

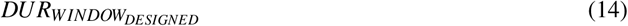

and the measured individual time window for a protocol window *m* as:-

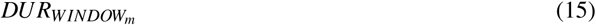

Similar to distance analogue accuracy calculations in the previous section we can calculate the measured time window mean and variance. There is no need for a scale factor or difference calculation to be applied here as the duration is measured in seconds no matter the protocol implementation.

The measured time window should tend to zero, as should its mean, thus:-

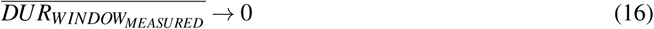

and variance, thus:-

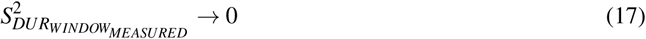

## 9 Longevity measures

Longevity concerns itself with the ability for a protocol to continue to detect and range other phones over a very long contact event period. This is useful for office, other work travel, and home based situations in order to accurately provide measurements during a full day of activity. It also represents a day’s worth of family activity in a crowded tourist location, or educational establishment.

Longevity can be thought of as the difference between continuity scores in the first hour of a work day compared with the last hour of the workday. I.e. the variation that may occur due to hardware or software flaws during a full day of operation of any proximity detection protocol.

In an ideal protocol there would be a very low (ideally 0) difference between the first and final hours of a day.

Thus by leaving a test running for 8 hours or more with a set of phones in the same location, the continuity scores can be used to calculate a longevity measure.

Recall from the continuity section 6.5 that the buckets missed score is represented as:-

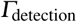

So our longevity measure can be represented as:-

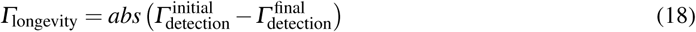

Where the initial measure is a delta score for the continuity sections for the first of 8 hours (120 readings using 30 second intervals) and the final measure is a delta score for the continuity sections for the last of 8 hours (120 readings again).

Note we are taking the absolute value as for a well performing protocol it is possible to have a better performing score in the final hour than the first hour. We should not give the protocol author additional credit here, as this still means there is variation over time which we must account for in order for our scoring to be fair.

This gives an overall percentage value of longevity which can be incorporated in to the probability of detection calculation.

## 10 Measures of detection

Continuity as discussed earlier is a measure of two devices that are in communication (either one-way or two-way) during a contact event. It is necessary though to come up with a measure that shows whether a device has seen another device during testing at all. Without this occurring then no continuity can be calculated as in a test you cannot ‘see’ that you’ve missed another phone in the recorded data. If we only considered continuity of detected devices then the overall protocol efficacy score would be over optimistic. There are two parts to detection:-

1. Whether device A can communicate with device B
2. Whether device A can identify device B

Only with both of the above being true can detection be said to have occurred. Thus it is not enough for two phones to be, for example, in communication over Bluetooth - they must also read the proximity detection identity payload correctly for a *detection* to have occurred.

The simplest measure for this is how many phones within a contact event test period saw any of the other phones. For ten phones in a test you would expect to see 90 pairwise connections (phone 1 seeing phones 2-10 is 9 detections, multiplied by 10 phones in the test).

Tested detection can therefore be defined as:-

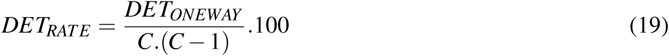

Where *DET*_*RATE*_ is the detection rate (a percentage), *DET*_*ONEWAY*_ is Phone A seeing Phone B (one-way detection and identification) for all phones, and C is the count of phones in the test.

Ideally for a fair test the top 10, or top 20, phones in use in a population of interest should be part of the test to arrive at detection rate. In the UK the top ten most popular phones account for over 15.02% of the population, the top 20 phones account for over 24.46% of the UK population [20].

We also recommend testing on older less popular phones so as not to disadvantage poorer or more marginalised sections of a population.

## 11 Measures of population reach

Before considering the overall performance of a protocol it is first necessary to determine the maximum numbers of devices this protocol will be able to run on. If a protocol is not capable of performing all of its functions on a particular model of device then it cannot hope to provide detection for those devices’ owners, or the people they come in to contact with.

### 11.1 Smartphone using population

Before a particular communications protocol is discussed it’s worth noting that not every resident of a country will have a mobile device. Older people and young children will not, although their movements are known by those that care for them and so can be accounted for by manual contact tracing. The highly mobile working age population, especially those whose companies issue work phones and who travel for work, are a key set of individuals to reach in order to control virus spread.

A study by the Office for National Statistics (ONS) in 2019 [9] shows the UK population was 66,435,550 of which 53,811,371 people were aged 16+, 4,571,627 were aged 10-15, and 8,052,552 were aged 0-9. We shall use these figures for ‘current’ UK population numbers.

A separate 2020 OFCOM study [10] shows smartphone ownership in the UK by age group was 82.49% for 16+, 66.90% for ages 10-15, and 9.11% for ages 0-9. We shall use these figures for ‘current’ UK phone ownership numbers.

The total percentage of the UK population (all ages) who own a smartphone is therefore ((82.49% × 53,811,371) + (66.90% × 4,571,627) + (9.11% × 8,052,552)) / 66,435,550 = 72.53%.

For the UK, therefore, this number is 72.53%. This figure shall be used for all calculations in this paper.

### 11.2 Protocol hardware support

Whilst the popular notion is that Bluetooth has been everywhere for years, in reality the support for parts of this standard varies greatly even amongst modern phone manufacturers. It is necessary to consider the range of this support. In order to discuss this in the specific rather than abstract we shall discuss Bluetooth Low Energy (BLE). BLE has been supported in the Bluetooth specification since Bluetooth 4.0 in June 2010. The current version is Bluetooth Low Energy 5.2 [26]. There are two key functions within the Bluetooth specification:-

– *Advertising*, or acting as a BLE Peripheral - A device advertising its presence and capabilities, allowing it to be *discovered* by a BLE scanning device, have its distance estimated, and optionally connected to
– *Scanning*, or acting as a BLE Central - A device looking for advertisements, reading that information, and optionally choosing to connect to and interact with a BLE device

It is worth noting briefly why BLE is supported on Mobile Phones. BLE is primarily used for the phone acting as a central to detect and connect to accessories such as Bluetooth headsets, external speakers, or your car’s audio system. Thus mobile phones must provide scanning in order to be useful in these consumer applications. Advertising - i.e. acting as a peripheral - is not a part of these activities. Not every phone therefore supports advertising even if they claim to be BLE compatible.

This means that a protocol that requires advertising will either have to not support some devices in the population, or to create a workaround to this gap in their support. It is worth noting that for many countries this will mean a purely advertising based protocol cannot service a significant amount of the population. [20, 21]

### 11.3 Protocol software support

Similarly, a protocol that uses features of an Operating System not available until a particular version will result in some older phones, which cannot be upgraded to that OS version, not being supported. This again can result in a significant proportion of the population not being supported. A protocol or contact tracing app that uses, for example, Elliptic Curve Encryption is only supported by iOS 10.7 and above. Whilst a small delta in coverage it must be considered.

### 11.4 Societal exclusion from protection

It’s worth noting that it’s normally older people or poorer families that can only afford basic models of phones, or to not upgrade to the latest phones with full Bluetooth support. There is, therefore, an information society and exclusion problem for proximity detection protocols. This is for government policy to address rather than a technical paper. It is morally incumbent upon all technologists to consider this impact though, and so we mention it here.

### 11.5 Measuring population reach

All of the previous measures in this section can be determined before a protocol is placed under formal practical testing. The other measures in this paper determine the efficacy of a protocol under testing. That efficacy should therefore also be factored in to any population reach calculations alongside the above effect. Thus we have two measures:-

1. Specification population effects - The maximum population reach for a particular protocol based on how it works in theory
2. Tested population effects - How continuity, completeness, and longevity affect the number of other devices seen and identified in real testing

The above are purely concerned with population reach and not epidemiological control. That is not the subject of this paper, but epidemiological control effect will be a function of the measures in this paper.

We propose that a simple percentage reach score made up of two parts shall be used, as follows:-

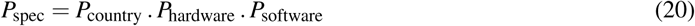

Where *P*_*SPEC*_ is the specification population effects, *P*_*COUNTRY*_ is the percentage of the country’s population who possess a device (Approximately 72.53% in the UK), *P*_*HARDWARE*_ is the percentage of that hardware in use that can be both detect and range (two-way detection and ranging) other devices, and *P*_*SOFTWARE*_ is the percentage of those devices in use that meet the requisite minimum software API and operating system functionality requirements in order to run that protocol.

Tested population effects shall be defined as:-

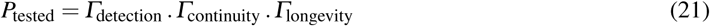

Where *DET*_*RATE*_ is the detection rate percentage, and *δ f lag*_*BUCKETS*_ is the missed buckets error rate percentage (i.e. how likely it is that in a detected device event within a particular 30 second distance estimation bucket is missed). See 6.5 for how this is calculated. Longevity is the changes of performance in continuity and detection over time during which a protocol is running, and potentially degrading during a day of use. See 18 for how this is calculated.

## 12 Overall efficacy of Contact Tracing

As per the 2004 Fraser et al paper [7] the efficacy of tracing value *εT* is calculated as “The efficacy of contact tracing (the y axis of Fig. 3) is the square of the proportion of the population using the app, multiplied by the probability of the app detecting infectious contacts, multiplied by the fractional reduction in infectiousness resulting from being notified as a contact.”. This formula is shown below:-

**Fig. 3.**
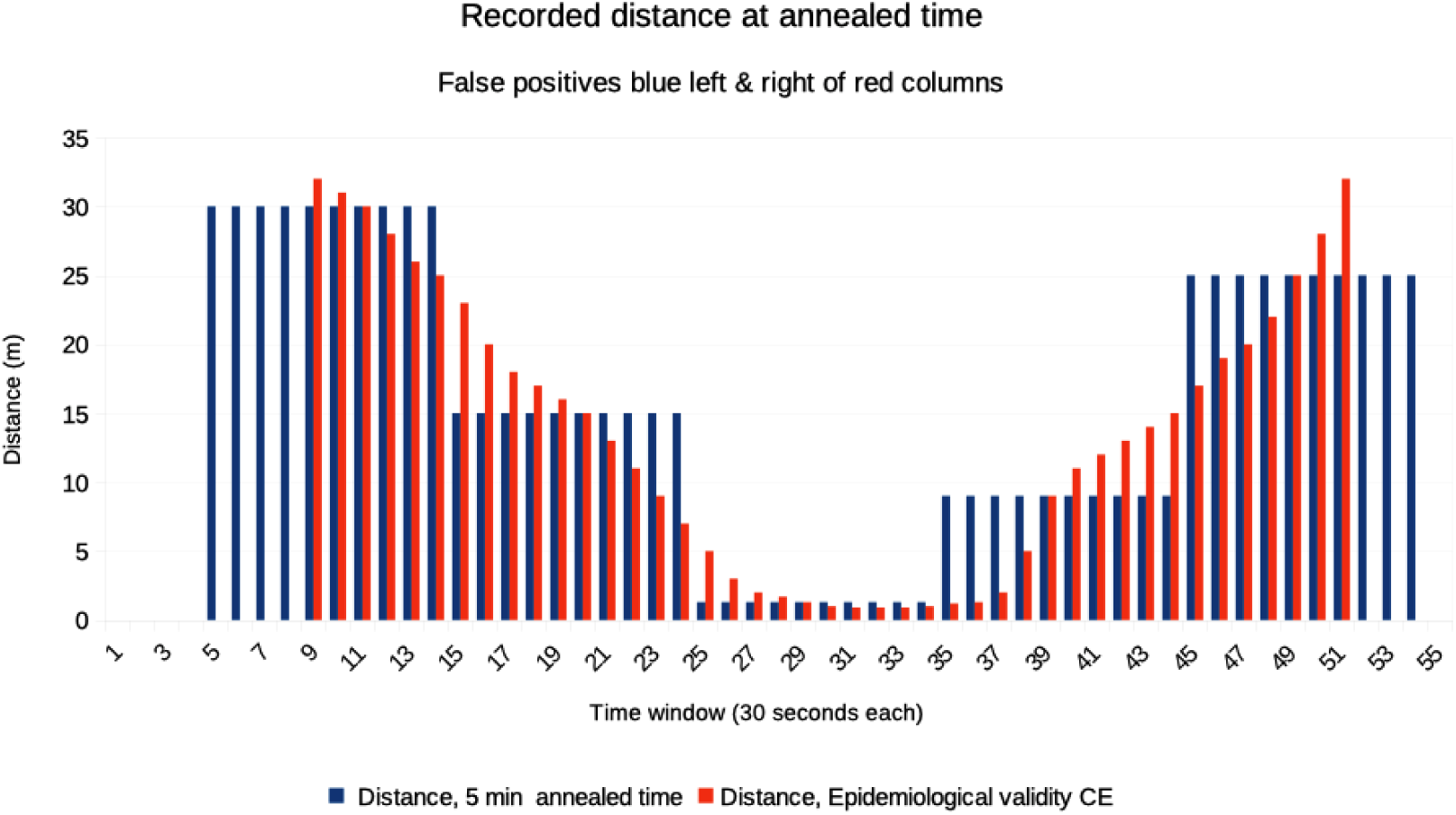
Distance analogue recordings for idealised versus 5-minute annealed protocols

**Fig. 4.**
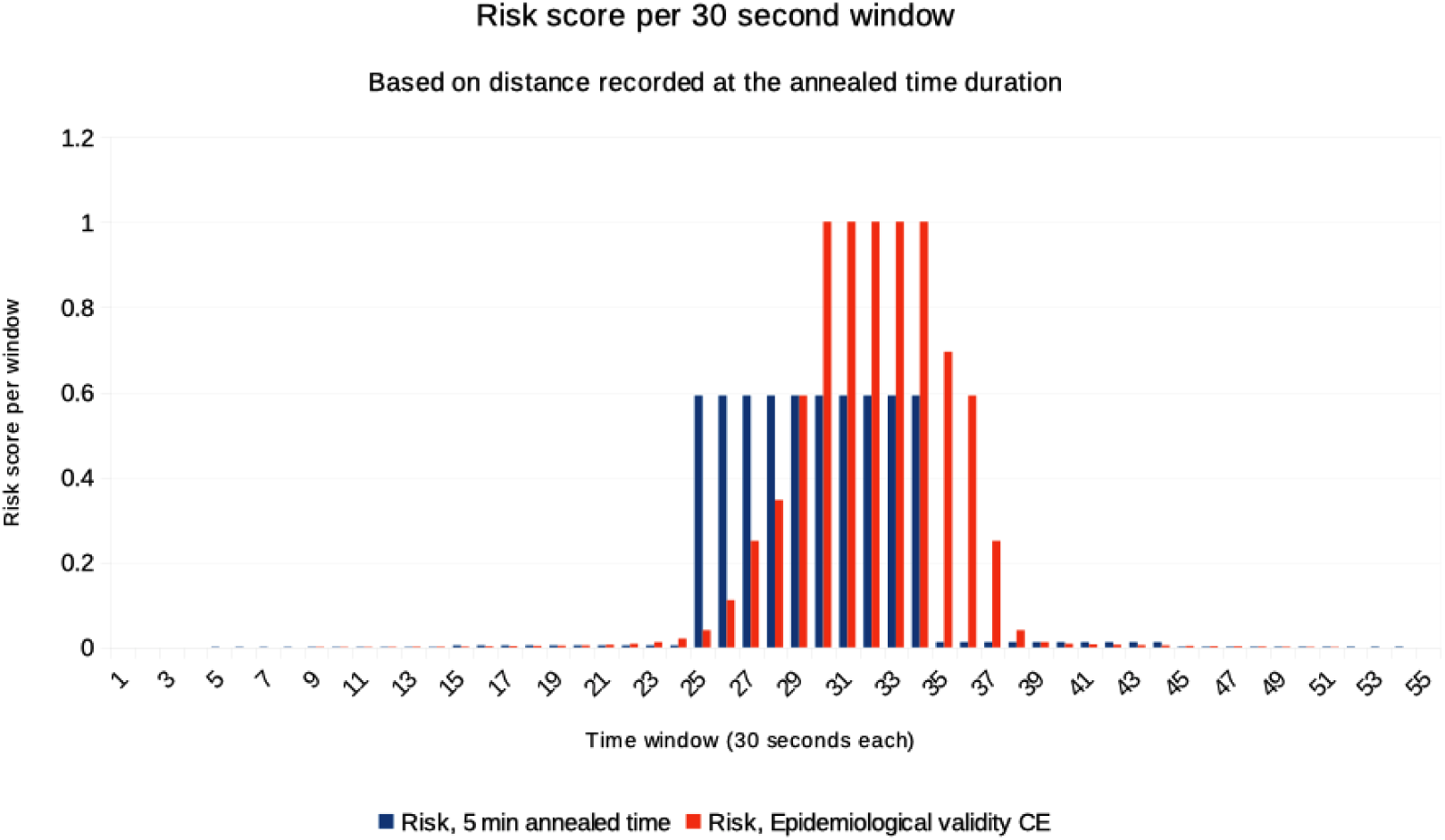
Risk recorded by idealised and 5-minute annealed protocols

**Fig. 5.**
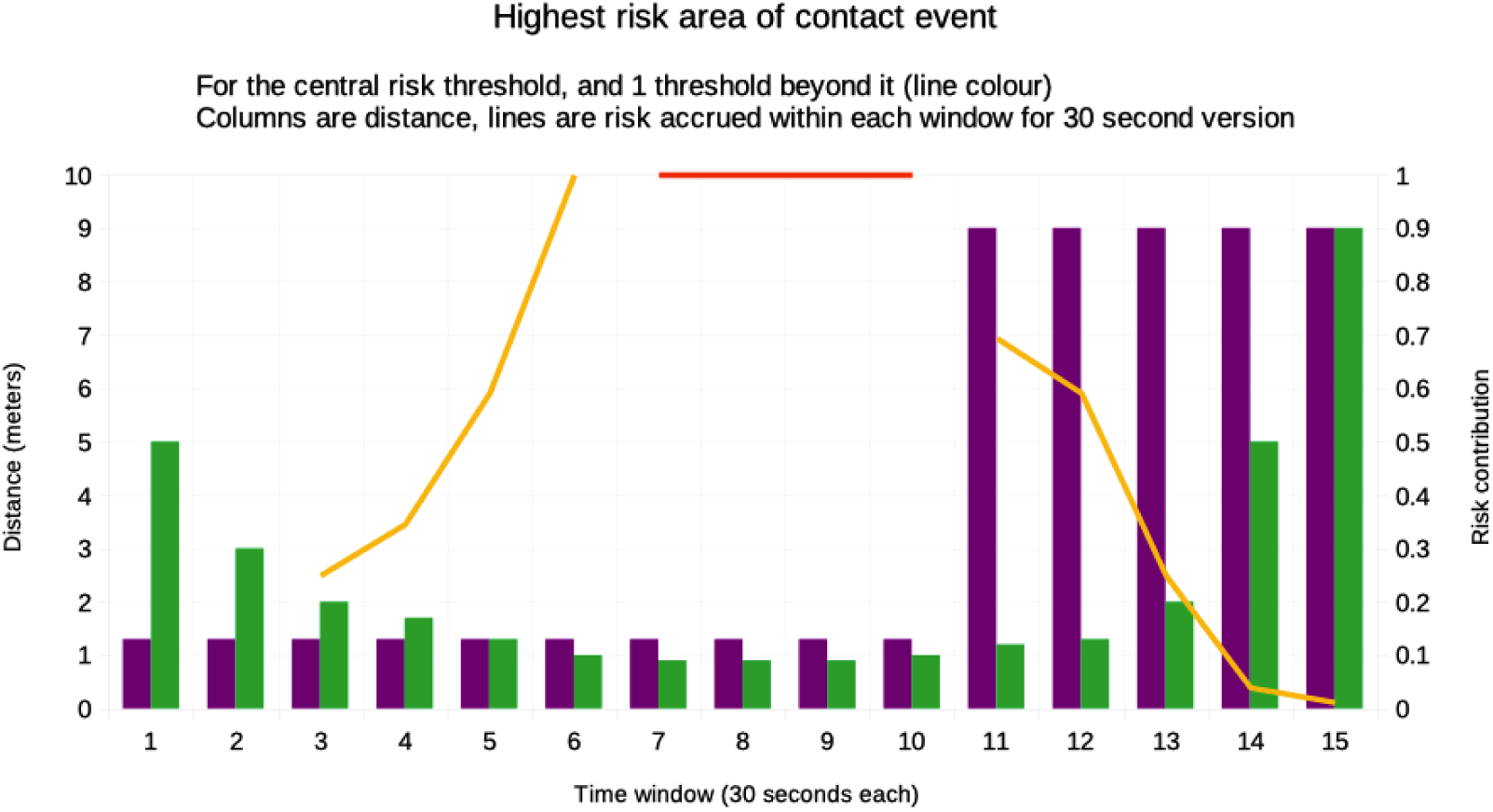
Risk threshold 1 and 2 around the nearest distance of a contact event. Green bars are exact risk measurement at 30 second intervals, purple bars are 5-minute interval measurements. The lines represent each time the contact accrued a risk exposure equal to the risk of 15 minutes at 2 metres, centred around the riskiest (nearest) part of a contact event.

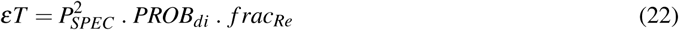

Where *f rac*_*Re*_ is the fraction of transmissions that are environmentally remediated (i.e. the control effect of asking people to self isolate with an app), which is 0.9 (i.e. 90% control effectiveness) in the Fraser et al paper [7]. So in the above we multiply by 90% for *f rac*_*Re*_.

Which is the protocol efficacy which includes continuity population reach, device hardware and software support, and longevity of contact tracing during a full work day, accuracy of distance estimation, and the error rate of the most risky period of each contact. We only include the most risky period so as not to skew the results towards more common contacts that would accrue much less risk to a person.

In our formulae in this paper the population using the app is the potential population, *P*_*SPEC*_. This was defined earlier. We now define the probability of detection itself, derived from the measures in this paper. The probability of the app detecting infectious contacts is therefore shown by the below:-

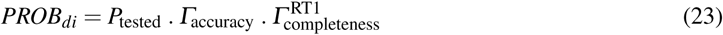

And expanded to:-

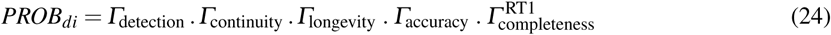

Ideally with this approaching 100%:-

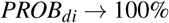

Expanding further to create the final efficacy of contact tracing measure from the Ferretti et al paper [6], gives:-

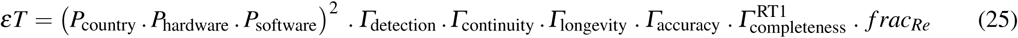

Applying the UK’s prevalence of mobile phones with Bluetooth Low Energy support, and assuming perfect support for phone operating systems in use, and perfect detection conditions between phones, the maximum UK effectiveness score that is possible is:-

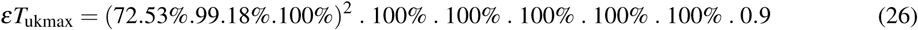

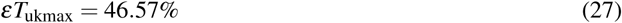

The above formula therefore provides the final measure which can then be used with the Fraser et al paper and the Ferretti et al paper to plot a given protocols’ contact tracing efficacy score and determine its controlling effect on a given infection’s spread.

Assuming a 90% effective isolation mechanism for ill people, the Ferretti et al paper [6] requires an estimated protocol efficacy of 30%, with the margin of error at the time of that paper’s publication being between 10% efficacy to 60% efficacy. This is required in order to slow the spread of COVID-19 (Fig 3 in that paper). It should be noted that the r=0 line on the chart shows a 30% protocol efficacy and a 90% efficacy in isolation will be enough to halt the spread of COVID-19 (an r=0 per day) for instantaneous contact tracing, assuming no error in the predictive model used.

Thus we should aim to find a protocol with at least a 30% efficacy rating by the above formula, but ideally with a 57% efficacy rating in order to cater for the uncertainty in the model. We shall therefore say that a protocol with 10% or more to have *limited effectiveness*, 30% or more to be *effective*, and 57% or more to be *very effective* in slowing COVID-19 spread. This is to enable layman’s interpretation of this numeric result. A simplified summary of the contributing factors to this formula is also available on our website. [14]

## 13 Fair formal testing methods

We have developed a set of formal test procedures that others can use to test their protocols effectively. Whilst out of the scope of this paper, we do detail these on our website. [15] This includes an ‘office test’ where ten phones are placed in two rows ideally 2m apart, simulating an office environment and providing detection, continuity, completeness, mean window length, and longevity measures. Also a Formal accuracy test to provide the accuracy measure.

## 14 Results

This section describes the results of our testing. We found that annealing time to a different window than which physically happened of 5 minutes reduces a probability of detection to around 20%, giving an efficacy score of just 4.46%. Doing this and bucketing the distance analogue using 10 buckets reduces these values further to 15.74% and 3.49% respectively. Even applying no buckets using a window value of just two minutes resulted in a poor score of 25.02% and 5.56% respectively.

We further applied the fair efficacy formula to a new protocol we developed called Herald. We found in real testing that Herald can achieve a probability of detection of 86.51%, giving a protocol efficacy score of 37.93%. It should be noted that this formal test included 10 phones with one of them having known performance problems. The theoretical maximum efficacy for any BLE based protocol in the UK is 46.57% given the prevalence of mobile phones in the UK. Herald therefore provides a very high probability of detection, exceeding the mean estimated score required by the Oxford paper to provide epidemiological control of COVID-19 in the UK population.

### 14.1 Measures for protocols in this paper

This section details the calculated formal comparison statistics for the three protocols described in this paper - the idealised 30 second window (Epidemiological) ideal variant, the two 5 minute annealed protocols - one that anneals just recorded contact time, (Annealed) another that anneals both contact time and buckets the distance analogue (Annealed & Bucketed) to ten RSSI buckets using a ceiling function, and a 2-minute window version without time annealing or RSSI bucketing. It is assumed both of the annealed protocols and the 2-minute protocol only use advertising to share identity information like some real-world protocols.

This results format also provides a useful sample *top sheet* layout in order to compare independent protocols’ formal assessment results.

As can be seen in Table 1, the completeness error for the simulated protocols is calculated by applying the 2 or 5 minute value to each 30 second interval. The error introduced is compensated for in the delta flag buckets (buckets missed) percentage.

**Table 1.**
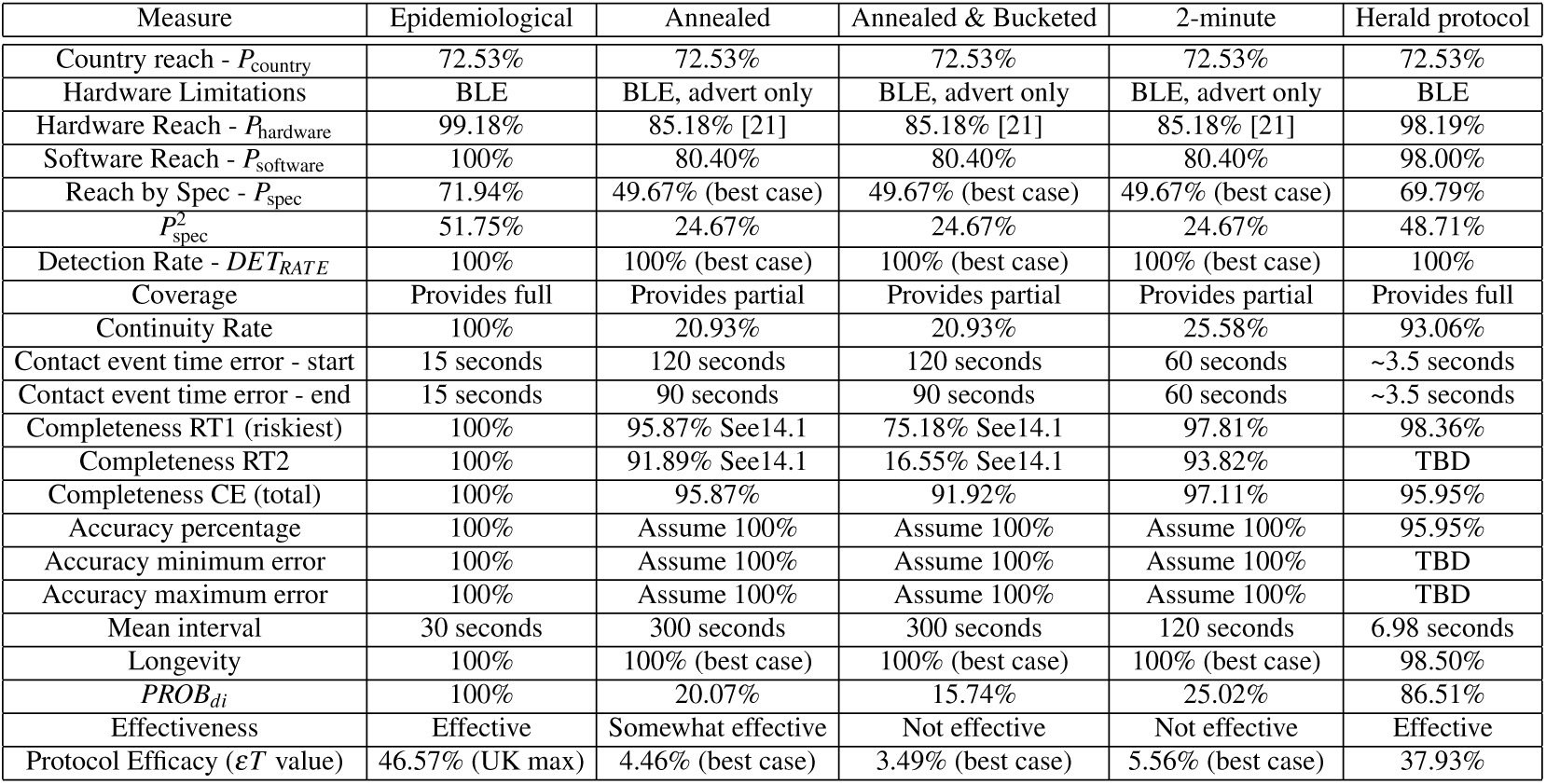
Simulated and tested performance of different protocols using the Fair Efficacy Formula

The above table’s errors were reverse calculated from distance in the original estimation source data using a simple algorithm for RSSI to distance conversion which is logarithmic and based on testing between an iPhone7 and iPhone7+. This formula was consistently applied for all simulated protocols.

As you can see from the first idealised protocol, the maximum efficacy score possible in the UK population is 46.57% due to mobile phone prevalence and mix.

Observe from the results that the second protocol that provides data only once every 5 minutes instead of once every 30 seconds and anneals that time to a specific time window as described in this paper can have an overall contact event risk completeness of 95.87%, which was also the completeness risk error during the riskiest part of the contact. This would lead to false positive and false negatives in real life.

The third protocol, Annealed and Bucketed, is the same as the second, except it only provides a bucketed distance analogue value rather than continuous decimal value. I chose to round up to the nearest 10 RSSI (I’m using positive RSSI only, not negative values as recorded), and thus an approximately 5% difference in bounded error. This 5% bounding error results in a near 25% risk score error in the riskiest (nearest) part of the sample contact event. This percentage can also become the percentage of over and under estimated risk scores, leading to a loss of confidence in the population if there are too many false positive alerts to test and self isolate.

Thus the third protocol can be said to over and under estimate risk by approximately 12.5% assuming an even spread of error, resulting in a potential 25% false positive or negative risk score. When multiplied across a range of contact events during a working day a protocol that both anneals time and buckets the distance analogue variable is likely to have a very high error rate when used for contact tracing, greatly limiting its effectiveness in use of such a protocol for limiting the spread of an outbreak. When you combine this with a continuity error (missed buckets) of over 79% there is a large potential for error in such protocols. This is true even though it is assumed in the above calculations to have a 100% detection rate - even though this may only be true for contact events lasting 5 minutes or more, or for protocols that only use advertising to exchange data. We also modeled a fourth protocol similar to the previous two, but instead measuring exact time from the start of a contact event (no time annealing), and with a shorter 2-minute window. This still had a high continuity error even though the completeness error was much reduced. This result shows that even assuming perfect conditions, taking existing advert-only protocols and reducing their time windows to around 2 minutes would still result in a much reduced efficacy score, and thus a lower control effect in practice.

This paper makes no claim as to the efficacy or otherwise of any real-life protocols. National contact tracing teams are encouraged to perform their own testing using the methodology and formula set out in this paper.

It should also be noted that the above results assume ‘immediate’ passing on (i.e. within 4 hours) of exposure notifications when someone is positively diagnosed. Some protocols therefore result in an effective delay of 24-72 hours. This results in a much lower virus transmission control effect in reality even if their efficacy scores were much improved. This is because potentially ill carriers would not be given advice to self isolate quickly enough, resulting in less disease spread control. See the Lorretti et al paper [6], Figure 3, for more details on this degradation.

We also quote efficacy results for our own Herald protocol that we developed from scratch after creating our efficacy formula. Using our formula allowed us to more quickly highlight and fix performance issues in development of the new protocol. We hope to collate test results for other protocols using our methodology on our website, if contributed by existing contact tracing teams worldwide. [16]

## 15 Discussion

In this section we discuss the limitations of our testing and formula, future work required, and implications of our work on contact tracing applications.

### 15.1 Conclusion

This paper provides a basis for fairly measuring and comparing distance estimation protocols without bias to a particular technology or method. When used with the Oxford Risk Model the formula presented should provide a consistent and realistic contact tracing efficacy score.

We now have a standardised way to measure the efficacy of proximity detection protocols used within contact tracing mobile apps. Governments can use this data to reassure citizens and encourage uptake of the apps. This will in turn increase the efficacy of contact tracing apps, and save lives, and prevent a return to national lockdowns. This work is also relevant to any outbreak of disease, not just COVID 19. This is especially true of the emergence of a new disease in future - keeping contact tracing apps available so they can be used at any time, perhaps with a general symptom reporting tool built in to spot new outbreaks or changes in diseases, is a good preventative steps governments can take to prevent future interference with normal life as has happened with COVID-19.

We found in the development of our own Herald protocol that the fair efficacy formula was effective in allowing us to create, test, and improve our own protocol from an efficacy score of 13% to one of over 40% within 5 weeks with a small development team. [18]

We have further shown that a wide range of efficacy score is possible between contact tracing applications, especially for those that use annealed time periods and bucketed distance analogue values.

Work should now be done on testing currently available proximity protocols and publishing the results publicly using this paper’s methodology. Only once this data is known can we begin to see how these protocols may work in practice to slow the spread of a future outbreak.

### 15.2 Literature comparison

We conclude that the efficacy formula presented in the Ferretti et al paper [6] can be calculated from real world testing of mobile contact tracing applications. We would encourage an epidemiological explanation of the use of 90% as the *f rac*_*Re*_ value as no basis for this value was presented in the paper. We discovered this value from the team’s code base on GitHub.

### 15.3 Limitations

Whilst the above formula provides a good approximation for real world efficacy for a contact tracing protocol there are of course going to be some potential improvements. The main one not discussed in this paper is the accuracy of the conversion mechanism from the distance analogue to a distance value. This paper’s formula assumes that, for a given pairing at a given RSSI reading, the conversion is perfect. Whilst we do include reading error in this paper - which is probably a larger contributor to error than mathematical conversion to distance - we do not consider the accuracy of the algorithm to convert the read distance analogue to an actual distance and its effect, if any, on risk attribution accuracy and thus efficacy. For protocols depending upon an integer RSSI value this would be useful to consider in future. As mentioned in a previous section, a single formula for distance conversion should not be relied upon given how RSSI is scaled in different Bluetooth chipsets. See section 8.2 for details.

This paper also does not consider physical location of phones on people’s person. I.e. front pocket vs. trouser pocket vs. in hand vs. in a purse. To fixate on such imperfections would be to deny any assistance to contact tracing through technology because you cannot make it perfect. It is the authors assertion that a control effect will never be perfect, just as medications’ efficacy can never be perfect, and thus it is not worth throwing the baby out with the bathwater or making a good protocol the enemy of the idealised perfect protocol described in this paper.

This author could not access the entitlement keys required to test these protocols and include any conclusions in this paper. Access to these are generally restricted to national governments using these protocols. The usefulness of the fair efficacy formula would be more compelling if we were able to add results from national governments’ own contact tracing apps. We would encourage contact tracing app teams to test their protocols and publish their results publicly.

Our own Herald protocol testing was limited by the fact we could not access many of the phone models that are popular in the UK. Our testing did not use the top ten phones. One advantage of this though is that we found and worked around a lot of Android and iOS Bluetooth issues that we would not have found by concentrating on the latest, most popular phones. Our continuity measure in formal testing was also made worse by the inclusion of a device (Pixel 3XL) that performed noticeably less well than the other phones in our test. Our efficacy score could therefore be higher than quoted in this paper.

### 15.4 Oxford Simulations

This paper?s efficacy results can be directly applied to the Oxford Big Data Institute (BDI) virus spread simulator, called OpenABM-COVID19 [27] enabling the evaluation of contact tracing protocols from an epidemiology perspective, e.g. predicting impact on infection and death rates, as well as demand on hospital beds.

The ‘Efficacy score’ (*εT* value) from this paper equates to the ‘traceable_interaction_fraction’ simulation parameter with two small modifications, removal of *Pcountry* and *Frac*_*RE*_. This gives:-

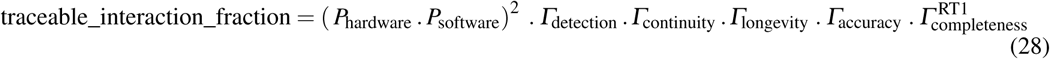

### 15.5 Future work

Now a low-level protocol that works across a large range of devices exists in the Herald protocol, the author aims to suggest a payload to transfer over this protocol that allows for its use in either a centralised or decentralised contact tracing application. This will provide international interoperability whilst allowing local jurisdictions to tailor their approach to one acceptable by its residents.

### 15.6 Implications of our work

We have shown that a standardised fair efficacy formula can be used to both simulate existing protocols’ efficacy but also to test protocols in real life. We have further shown that it can be used to better direct development of a protocol. This implies that existing contact tracing development teams can use our fair efficacy measure alone to quickly measure and improve the effectiveness of their own protocols.

Our results along with the Ferretti et al paper [6] show that an effective protocol can control the spread of COVID-19 if used in an app with same day exposure notification.

We have further shown that protocols which bucket their distance analogue or have time windows larger than 30 seconds will have a lower efficacy score, and therefore also a lower control effect on the spread of a disease such as COVID-19.

## Supporting information

Fair Efficacy Formula calculations

## Data Availability

Data is available as part of the submission. Further related research can be found on https://vmware.github.io/herald

